# Effect of covid-19 lockdown on child protection medical assessments: a retrospective observational study in Birmingham, UK

**DOI:** 10.1101/2020.08.09.20170977

**Authors:** Joanna Garstang, Geoff Debelle, Indu Anand, Jane Armstrong, Emily Botcher, Helen Chaplin, Nutmeg Hallett, Clare Morgans, Malcolm Price, Ern Ern Henna Tan, Emily Tudor, Julie Taylor

**Affiliations:** Birmingham Community Healthcare NHS Foundation Trust; College of Medicine and Dentistry, University of Birmingham, Edgbaston, Birmingham, UK; Birmingham Women’s and Children’s NHS Foundation Trust; Birmingham and Solihull Clinical Commissioning Group

**Keywords:** child abuse, non-accidental injury, paediatrics, child protection medical examination

## Abstract

**Objectives:** To determine any change in referral patterns and outcomes in children (0-18) referred for child protection medical examination (CPME) during the covid-19 pandemic compared to previous years.

**Design:** Retrospective observational study, analysing routinely collected clinical data from CPME reports in a rapid response to the pandemic lockdown.

**Setting:** Birmingham Community Healthcare NHS Trust, which provides all routine CPME for Birmingham, England, population 1.1 million including 288,000 children.

**Participants:** Children aged under 18 years attending CPME during an 18 week period from late February to late June during the years 2018, 2019, and 2020.

**Main Outcome Measures:** Numbers of referrals, source of disclosure and outcomes from CPME

**Results:** There were 78 CPME referrals in 2018, 75 in 2019 and 47 in 2020, this was a 39.7% (95%CI 12.4-59.0) reduction in referrals from 2018 to 2020, and a 37.3% (95%CI 8.6-57.4) reduction from 2019 to 2020. There were fewer CPME referrals initiated by school staff in 2020, 12(26%) compared to 36 (47%) and 38 (52%) in 2018 and 2019 respectively. In all years 75.9% of children were known to social care prior to CPME, and 94% of CPME concluded that there were significant safeguarding concerns.

**Conclusions:** School closure due to covid-19 may have harmed children as child abuse has remained hidden. There needs to be either mandatory attendance at schools in future or viable alternatives found. There may be a significant increase in safeguarding referrals when schools fully re-open as children disclose the abuse they have experienced at home.

**Article summary: Strengths and Limitations of the Study:**

- This is a highly robust study: we obtained CPME reports for 97% of CPME referrals during the study period.
- We ensured consistency of data extraction by double reviewing every report, with further consensus discussions for the few cases that raised uncertainties.
- The team extracting the data comprised highly experienced paediatricians with expertise in child abuse.
- One weakness is that we only considered minor injuries from outpatient CPME, excluding those admitted to hospital, so our findings do not include those with more serious NAI, however they would be taken to hospital for treatment due to the severity of their injuries.

## Introduction

Nearly 400,000 children in England each year are defined as ‘children in need’; these are children who require additional services, including safeguarding, to maintain a satisfactory level of health or development(1). Since the lockdown began, there are burgeoning concerns that child protection referrals have decreased, with professionals reporting limited opportunities to make accurate assessments of children’s needs(2). Statutory guidance sets out the specific roles and responsibilities of agencies for undertaking child protection enquiries when a child or young person is referred for suspected maltreatment(3), including formal child protection medical examinations (CPME). The purpose of CPME is to provide a holistic assessment of the child’s health, document any injuries and determine possible causes including the reasonable likelihood of injuries being inflicted or non-inflicted. A report is provided to inform any child protection investigations. CPMEs are performed or supervised by an experienced consultant paediatrician (4), adhering to rigorous standards in respect of consent; conduct of the examination; documentation of history; findings and formulation; photo-documentation; and report writing(5), with reports subject to regular peer review(6).

Birmingham is the second largest city in the UK, with a diverse population and is the largest local authority in Europe. It is also a relatively young city, with 23% of its population being children under the age of 16 years(7). The proportion of children subject to a child protection plan is higher than for the UK as a whole(8) and thirty-five percent of children live in poverty(8). In Birmingham CPMEs are generally undertaken within a community setting during working hours, often for children who have disclosed maltreatment to school or nursery staff, who then refer them to Birmingham Children’s Trust (social care). Children with suspected sexual abuse are assessed separately, within specialised, regional child sexual assault referral centres. Hospital-based paediatricians perform CPMEs for those children with more significant injuries requiring treatment and for out-of-hours referrals. During the covid-19 lockdown the community based CPME service provided extended hours (6 April to 23 May 2020) that covered evenings and weekends to minimise hospital attendance so an increase in referrals for CPME was expected.

Schools are at the frontline of child safeguarding; educational staff are often the first to report potential child abuse. This raises concerns that vulnerable children are now invisible to professionals and potentially ‘at risk’ in homes where families face even greater hardships(8). Such ‘collateral damage’(9) has been borne out by evidence that only 10% of children on a child protection plan or ‘in-need’ were attending schools that were remaining open specifically for their benefit and even where schools are open for selected year groups, attendance remains very low (10).

Although there has been much professional concern about the potential risk children have faced at home there have been limited data, with one report of an increase in abusive head trauma noted in London (11) and a short report from the North-East of England noting a dramatic decrease in CPME referrals (12). This current study was designed as a rapid response to fill gaps in knowledge about child protection referrals during the covid-19 pandemic.

The aim was to determine differences in the number and outcomes of child protection referrals for CPME in Birmingham during the covid-19 pandemic lockdown (March to June 2020) compared with the same periods in 2018 and 2019. Our research questions were:

What is the difference in child protection referrals during the covid-19 pandemic compared to previous years?

Are there differences in demographic details, referral source and outcomes for children presenting for child protection medical examination during the covid-19 pandemic compared to previous years?

## Methods

### Study design

Retrospective observational study of referrals for CPME. It adhered to STROBE guidance (13).

### Setting and sample

All children aged 0-18 attending for CPME at Birmingham Community Healthcare Trust (BCHT), England. BCHT provides specialist CPME for the population of Birmingham, total population 1.1 million of which 288,000 are children aged <18 (7). Data were collected for all CPME for 18-week periods in 2018, 2019 and 2020, from the last week in February, when schools returned following the half-term holiday, to the end of June.

### Procedure

We obtained a list of all children referred for CPME from the booking service and accessed the electronic patient records (EPR) for these children, obtaining copies of reports from CPME. We read the reports, and completed an anonymised data extraction form for each CPME (on-line supplementary file 1). The data collection form was in three parts: i) child demographic data, including age, gender, school age group (pre-school, primary, secondary, post-16), in a special school or not, ii) referral details including whether an index case or referred as a sibling group, source of initial disclosure, who the allegation was against, whether the child had previous referral to social care and if so, the current social care status, and whether the child had ever been on a child protection plan, and iii) outcomes of the CPME including whether there were physical findings to support non-accidental injury (NAI) or neglect, and, if so, what were the physical findings; likelihood of NAI; whether NAIs were present on more than one body part; were their injuries consistent with previous NAI; whether the report indicated significant safeguarding concerns; if the concerns were related to factors other than NAI; and, if so, what?

Outcomes were taken either directly from the conclusion of the CPME report, or if the conclusion was unclear, were determined based on the description of injuries and events within the report. If the CPME was not available, we used the EPR for demographics, referral source and safeguarding history, omitting data on outcomes of CPME.

Prior to commencing data extraction, all the clinicians reviewed 10 anonymised CPME reports which were then reviewed and discussed by the whole group. This enabled any differences in interpretation of CPME to be resolved and ensured quality and consistency of data extraction.

Clinicians worked in pairs, consisting of a specialist consultant in child protection (either Named or Designated Doctors for Safeguarding) and a specialist trainee in paediatrics, all of whom have a minimum of four years postgraduate medical training in child health. Each case had data extracted independently by the consultant and trainee, to ensure consistency. In the event of disagreement the case was reviewed by another consultant.

### Study size

As this was an observational study, no sample size calculation was undertaken. The time period included the last week in February which was before there was significant concern in schools about covid-19. Data collection continued for the month of June to enable any change in referral CPME patterns with the partial reopening of some primary schools.

### Statistical analysis

Anonymised data were entered into SPSS. Cases were analysed by the year of referral. If children had more than one CPME during the study period, each CPME was considered as a separate case. Referral rates between years for the whole 18-week period were compared using incidence rate ratios (IRR). IRRs for two weekly time-periods comparing 2018/19 with 2020 were also calculated and plotted on a graph with 95% confidence intervals. To compare differences in variables between the years, Kruskal-Wallis tests were run for continuous variables (age, number of types of injuries) and chi-square tests were run for categorical variables.

### Ethics

This study involved clinicians analysing routinely collected patient data, from patients within their own clinical service, it therefore does not require HRA ethical approval. The study was approved by the Research and Innovation Department in Birmingham Community Healthcare Trust.

### Patient and public involvement

As a rapid observational study using retrospective records we were unable to include children who had been through a CPME or their parents in the study. However we have a Children and Young People’s Advisory Group whom we intend to involve in the dissemination and guidelines for practitioners.

## Results

There were 200 CPMEs during the study period; 193 had CPME reports available with complete information from 191.

### Referral numbers

There were fewer CPME referrals in 2020 compared to previous years, as shown in figure 1. There was a 39.7% (95%CI 12.4-59.0) reduction in referrals from 2018 to 2020, and a 37.3% (95%CI 8.6-57.4) reduction from 2019 to 2020. The IRR for 2020 compared to 2018/19 was 0.61 (95%CI 0.43-0.86) showing an overall reduction of 39% (95% CI 14%-57%).

**Figure 1.**
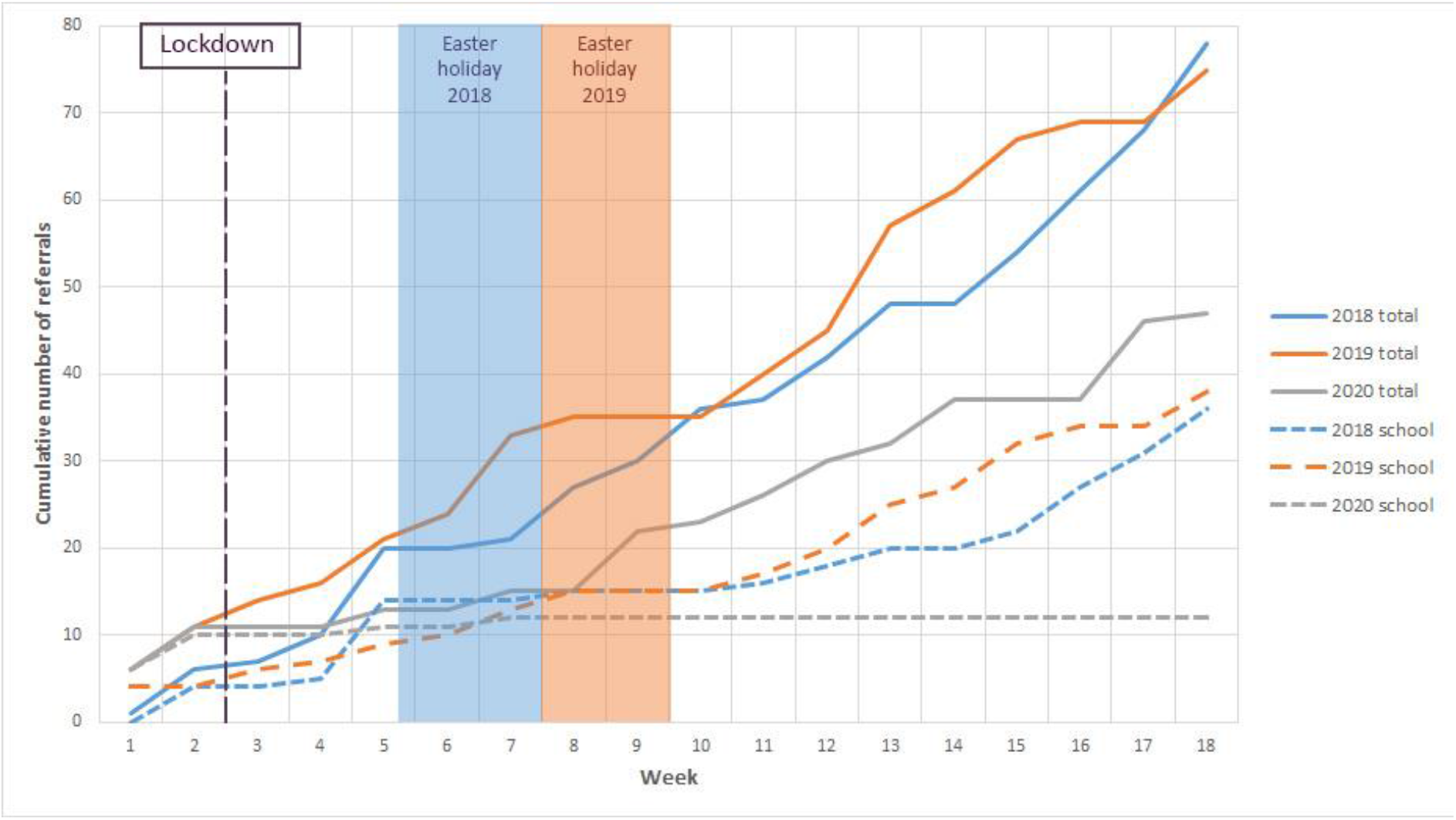
Cumulative number of referrals

The two weekly data shows that there was a significant drop in referrals for a 6-week period from weeks 3/4 to weeks 7/8, see figure 2. There was some evidence of an increase in referrals during weeks 9/10 in 2020 after which referral rates were broadly similar, with all confidence intervals crossing 1, apart from weeks 15/16 when there were no referrals in 2020.

**Figure 2.**
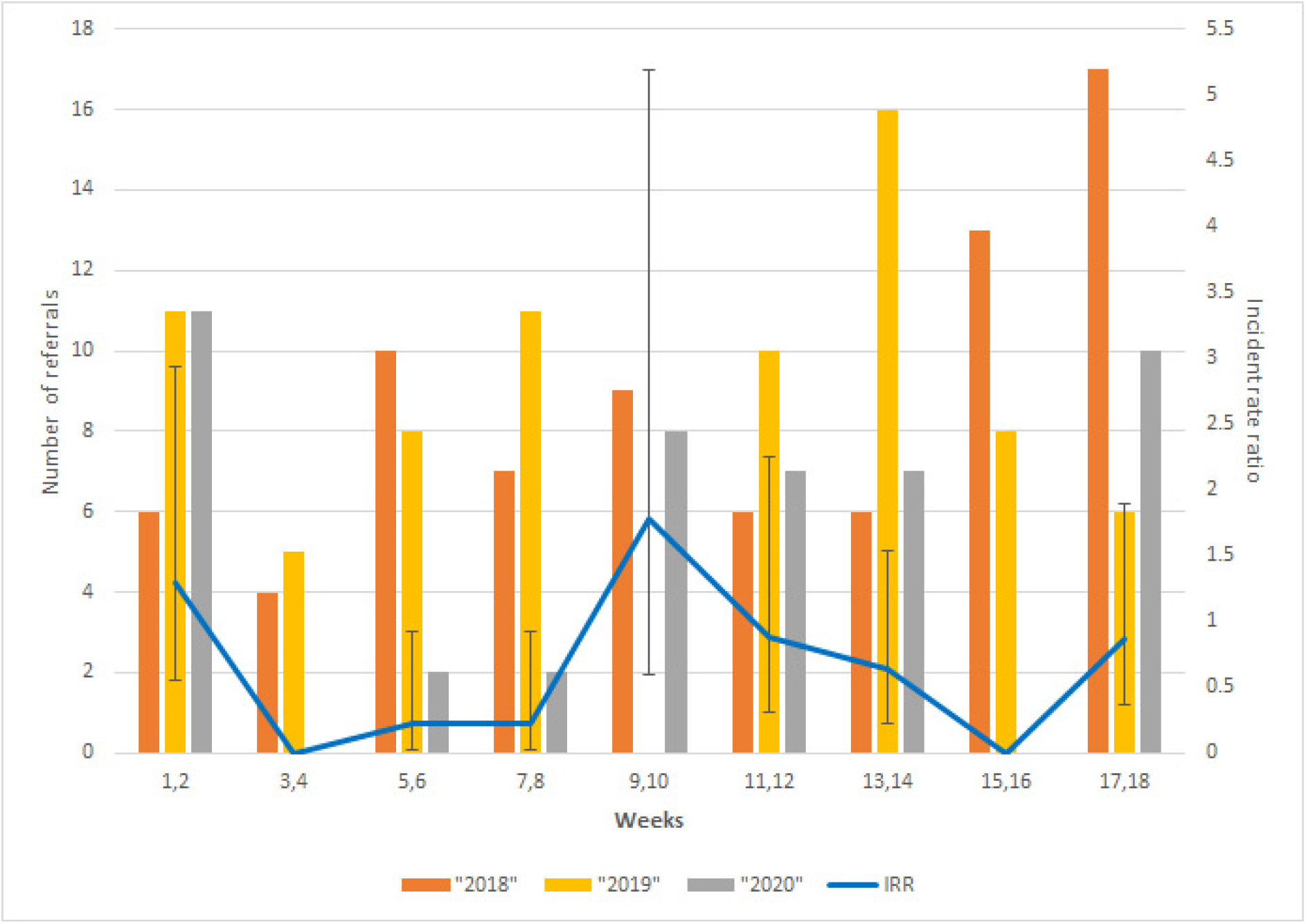
IRR and totals of weekly referrals

### Secondary Outcomes

A summary of referrals, demographics, social care history and outcomes of CPME is shown in table 1.

**Table 1.** Summary of key findings

|  | All | 2018 | 2019 | 2020 | p |
| --- | --- | --- | --- | --- | --- |
| **Sample N** | 200 | 78 | 75 | 47 |  |
| **Age in months Median (IQR)** | 69 (85) | 72.5 (76) | 70 (109) | 55 (77) | .598 |
| **Gender N Female (%)** | 73 (36.5) | 32 (41.0) | 31 (41.3) | 10 (21.3) | .046 |
| **School status N (%)** |  |  |  |  |  |
| Pre-school | 85 (42.5) | 34 (43.6) | 30 (40.0) | 21 (44.7) | .637 |
| Primary (Reception – Year 6) | 78 (39.0) | 32 (41.0) | 26 (34.7) | 20 (42.6) |  |
| Secondary (Year 7-11) | 32 (16.0) | 11 (14.1) | 16 (21.3) | 5 (10.6) |  |
| College / 6<sup>th</sup> form (Year 12-13) | 5 (2.5) | 1 (1.3) | 3 (4.0) | 1 (2.1) |  |
| **Is child an index case (vs sibling or household contact) N Yes (%)** | 134 (67) | 46 (59.0) | 56 (74.7) | 32 (68.1) | .117 |

| **Source of referral (who did child disclose abuse to, or who initiated CPME referral) *N (%)*** |  |  |  |  |  |
| --- | --- | --- | --- | --- | --- |
| School or Early Years staff | 86 (43.9) | 36 (47.4) | 38 (52.1) | 12 (25.5) | .015 |
| Social care staff | 22 (11.2) | 10 (13.2) | 3 (4.1) | 9 (19.1) |  |
| Police | 22 (11.2) | 11 (14.5) | 7 (9.6) | 4 (8.5) |  |
| Family member | 36 (18.4) | 9 (11.8) | 17 (23.3) | 10 (21.3) |  |
| Medical professional | 4 (2.0) | 2 (2.6) | 2 (2.7) | 0 (0.0) |  |
| Foster carer | 7 (3.6) | 1 (1.3) | 2 (2.7) | 4 (8.5) |  |
| Sibling current inpatient due to NAI | 12 (6.1) | 6 (7.9) | 1 (1.4) | 5 (10.6) |  |
| Other | 7 (3.6) | 1 (1.3) | 3 (4.1) | 3 (6.4) |  |
| **Was child known to social care prior to CPME referral? *N Yes (%)*** | 151 (75.9) | 58 (74.4) | 56 (75.7) | 37 (78.7) | .857 |
| **Is child an open case to social care now *N Yes (%)*** | 106 (53.3) | 39 (50.0) | 38 (51.4) | 29 (61.7) | .409 |
| **Are there physical findings to support NAI or neglect *N Yes (%)*** | 98 (51.0) | 32 (41.0) | 46 (59.0) | 27 (57.4) | .071 |
| **Does the report indicate significant safeguarding concerns? *N Yes %*** | 180 (93.8) | 69 (88.5) | 64 (95.5) | 47 (100) | .014 |

There were significantly fewer referrals made by school or early years staff in 2020 compared to other years, with only two school referrals received after lockdown. There was no increase in referrals or disclosures from other sources. In each year, several referrals were initiated when children disclosed abuse to grandparents and non-resident parents or by relatives who witnessed abuse.

There were significantly fewer girls referred in 2020. In total across all years, 67% of children were index cases who disclosed potential abuse, or had concerning injuries noted by others leading to referral; the remaining 33% were siblings of these index cases. Across all years 75.9% of children were known to social care at any time prior to CPME, 53% were open cases in receipt of support from social care immediately prior to CPME and 39% were currently or had previously been subject to a child protection plan (where maltreatment has been substantiated).

The findings in 51% of all CPME were that there was evidence of non-accidental injury (NAI) or neglect, with 55% of these children having injuries, typically bruising, on more than one area of their body implying more significant NAI. In 90% of all CPME it was concluded that there were significant safeguarding concerns: even if there was not evidence to substantiate NAI, there were significantly fewer children in this category in 2018, but the reasons for this are unclear. There was no other statistical evidence of differences in demographics, social care histories, referral sources and outcomes; further details are shown in on-line supplementary table 2.

## Discussion

### Statement of principal findings

This study found a significant drop of 39% (95%CI 14-57%) in CPME referrals during 2020 compared to previous years. This drop coincides with the near total absence of referrals made by schools after school closure in March, with no recovery in school referrals after schools partially re-opened in June. Referrals from other sources did not increase in 2020, showing that other agencies did not fully compensate for school closure. The children referred for CPME in 2020 had similar social care histories to other years with the majority being previously known to social care and approximately half being open cases at the time of referral. In all years, the vast majority of CPME reports concluded that there were significant safeguarding concerns relating to physical abuse, domestic violence, emotional abuse or neglect.

Our trust is the largest provider of community paediatric services in England, managing all requests for outpatient CPME for Birmingham residents. The extended hours offered during lockdown meant that we could include children with minor injuries needing CPME who ordinarily would be managed by acute hospital trusts, so our findings may actually be an under-estimation of the decreased referral rate. Our findings should be generalisable outside of Birmingham, as this is a large multi-cultural city with above average levels of social deprivation and is the largest Local Authority in Europe.

### Strengths and weaknesses in relation to other studies, discussing important differences in results

Although the drop in CPME referrals has been noted elsewhere in the UK (12), the longer duration of our study enabled us to examine any effects of the partial re-opening of schools. Our detailed analysis of referral details and outcomes identified the change in referral patterns this year, which is a novel finding. As our CPME service covers a fixed population, we can be certain that changes in referral patterns are genuine, unlike tertiary paediatric centres whose referrals are determined by clinical need not home address (11).

### Meaning of the study: possible explanations and implications for clinicians and policymakers

Our findings further evidence the hidden harm to children from covid-19. The significant decrease in CPME referrals is likely largely a result of school closure and the partial re-opening of schools has not altered this trend. Attending school provides children and young people with access to a trusted adult and a safe space outside of the family home. Removing this provision increases the potential risk of abuse going unseen. Many schools have made strenuous efforts to maintain contact through remote methods, but these are not always private and it is not known who else may be in the room. Although UK government guidance was for vulnerable children, identified as those with an allocated social worker, to continue attending school, less than 10% did so (10). Nearly half of those referred for CPME were not in this category so had no protection. Disclosures to school staff by older children also protects younger siblings from abuse. Missed sentinel NAI such as bruising, may lead to children subsequently presenting with serious injuries (14) (15). These sentinel NAI are typical of community CPME referrals and the drop in referral rate therefore represents a much greater risk of harm. While UK government policy is for mandatory school attendance from September, it is vital that this is encouraged and enforced by schools given that currently less than 40% of eligible primary school pupils are attending (10). Low attendance rates may enable abusing parents to keep their children at home with few questions asked: there must be robust face to face welfare checks for those who do not attend. Once back at school, many children may disclose abuse that occurred during closure, and children’s services may struggle to meet demand. As months will have passed since the abuse, there may be little physical evidence to support allegations, in turn reducing the weight of corroborative evidence to support child protection measures and risking children feeling they are not believed. Child abuse carries long-term risks for cumulative physical and mental health problems(16,17), and without intervention a cycle of intergenerational poor parenting and abuse may result(18). There were 30 fewer CPME referrals than expected during 2020: given that Birmingham accounts for 2% of children referred for social care assessment nationally (1) we estimate that there are approximately 1500 (95%CI 538-2192) potentially abused or neglected children in England who remain hidden from services. This number may be considerably greater with the suspected rise in rates of child abuse during lockdown. We face an epidemic of unreported, unrecognised child abuse with long-term implications for society as a whole. Getting all children back into school will reduce the risk, but may not undo the harm that has already occurred. Should there be a further lockdown, safeguards must be put in place to prevent vulnerable children coming to harm.

### Unanswered questions and future research

We need to continue to evaluate CPME referral patterns and outcomes as children return to school, to help understand the hidden harms from covid-19. There should be robust analyses of inpatient NAI cases to determine any increase in severe injuries. Research should include hearing children’s lived experiences so that appropriate safeguards can be put in place should schools have to close in future. Longer-term research is needed to ascertain and treat the mental health and behavioural outcomes that may result from abuse during school closures. As ‘child safeguarding is everyone’s business’(19) learning how to protect children during an event such as covid-19 should be a multi-agency process, and perhaps the National Safeguarding Children’s Panel should take this forward.

## Data Availability

Data are drawn from clinical child protection reports of individual children and cannot be shared. The disaggregated and anonymised abstraction files may be shared at reasonable request from the first author

## Data sharing statement

Data is drawn from clinical child protection reports of individual children and cannot be shared. The disaggregated and anonymised abstraction files may be shared at reasonable request from the first author.

## Author Contributions

JG conceived the idea, the protocol was designed by JG, GD, JT, JA, and IA. The data extraction tool was piloted, revised then used for data extraction by JG, GD,HC, JA, IA, ET, EET, CM, and EB. The data analysis was undertaken by JG, NH and MP. Drafts of the paper were written by JG, GD, NH and JT with all authors contributing. Information for the paper came from the clinical records for each child. JG is guarantor of the article.

Word Count: 2662

